# Assessment of Long-Lasting Insecticidal Net (LLIN) Ownership, Utilization, and Associated Barriers in Malaria-Endemic Communities of Ethiopia

**DOI:** 10.64898/2026.03.25.26349322

**Authors:** Abate Waldetensai, Geremew Tasew, Delenesaw Yewhalaw, Habte Takie, Bokretsion Gidey, Solomon Kinde, Fekadu Gemechu, Selam Yirga, Esayas Kinfe, Alemnesh Hailemariam, Henock Tadesse, Hiwot Solomon, Gudissa Assefa, Dereje Dilu, Seife Bashaye, Yonas Wuletaw, Bedri Abdulatif, Tilahun Kebede, Samson Tadiwos, Gashaw Gebrewold, Samuel Hailu, Fitsum Tesfaye, Getachew Tollera, Messay Hailu, Yan Guiyun, Araya Eukubay, Araya Gebresillassie

## Abstract

**Background:** Malaria remains a critical global health challenge, with over 68% of Ethiopia’s population living in at-risk areas. While Long-Lasting Insecticidal Nets (LLINs) are a cornerstone of prevention, their effectiveness depends on consistent use. This study aimed to assess LLIN ownership and utilization patterns and identify socio-behavioral and physical determinants of their effectiveness in endemic communities.

**Methods:** A community-based, cross-sectional survey was conducted from October 2024 to January 2025 across 11 administrative regions in Ethiopia. Using a two-stage stratified cluster sampling technique, data were collected from 9,222 households (34,427 individuals) through face-to-face interviews and direct physical observations. Data analysis was performed using the SPSS Complex Samples module and hierarchical multivariable logistic regression.

**Results:** The survey found a household LLIN ownership rate of 71.5%, while the proportion of sufficient LLINs for every two people was 58.3%. Among those who owned nets, the overall utilization rate was 59.9%, with significantly higher rates in rural areas (72.7%) than in urban areas. Vulnerable groups achieved higher usage levels, specifically pregnant women (78.5%) and children under five (67.2%). Multivariable analysis indicated that age and pregnancy status were the strongest predictors of LLIN use, with ORs of 0.258 (p < 0.001) and 0.662 (p < 0.001), respectively. Major barriers identified included a 60.5% lack of confidence in hanging nets (p < 0.001) and a widespread misconception (64.1%) that malaria risk is restricted to the rainy season.

**Conclusion:** Although Ethiopia has made strides in LLIN ownership and prioritized protection for vulnerable demographics, overall utilization remains below the 80% threshold required for community-wide protection. To bridge the gap between ownership and consistent use, national strategies should transition toward skill-based interventions and targeted communication to address practical barriers and seasonal misconceptions.

## Background

Malaria is one of the most significant and preeminent global health challenges, defined by the magnitude of its morbidity and the severity of its clinical outcomes [1]. Recent epidemiological data from 2022 estimated a global burden of 249 million cases and 608,000 deaths, of which approximately 93.6% of all cases and 95.4% is in Africa [2], following substantial reductions in disease burden between 2000 and 2015 [3]. High-transmission intensity is largely localized, with five African countries, Nigeria (25.9%), the Democratic Republic of the Congo (12.6%), Uganda (4.8%), Ethiopia (3.6%), and Mozambique (3.5%) collectively representing more than half of the global disease burden [4]. Children under five years of age remain the most severely impacted demographic [5], representing roughly 75% to 80% of all malaria deaths in Africa [1,6]. This high mortality is compounded by the physiological vulnerability of pregnant women, where malarial infection contributes significantly to maternal anemia, stillbirth, and low birthweight, impacting hundreds of thousands of infants annually [7]. The Global Technical Strategy (GTS) for Malaria 2016–2030 establishes a mandate to reduce case incidence and mortality rates by at least 90% by 2030 compared to 2015 levels [8]. However, the world is currently off track for these milestones; for instance, the 2024 global mortality rate was more than three times the target level [9]. LLINs are recognized by the World Health Organization (WHO) as one of the most cost-effective tools for reducing transmission and have been credited with averting nearly 70% of cases globally [10,11].

In Ethiopia, approximately 75% of the total landmass is considered malarious, placing roughly 68% to 69% of the population, approximately 52 to 60 million people, at risk of infection [7,12,13]. Malaria transmission is characterized as seasonal and unstable, occurring primarily in areas below 2,000 meters above sea level where it is heavily influenced by rainfall patterns and altitude [14,15]. Ethiopia’s diverse ecological landscapes provide a breeding habitat for the *Anopheles arebiensis*, the primary vector of malaria transmission in the country [16,17]. Environmental factors and climate change pose a substantial threat to control efforts, as rising temperatures potentially expand malaria risk zones into the highlands and exacerbate the instability of transmission [18,19]. Long-Lasting Insecticidal Nets (LLINs) and Indoor Residual Spraying (IRS) are the major cornerstones of the Vector control program as the mainstay interventions in Ethiopia’s malaria prevention strategy [13,20,21]. The WHO and the Ethiopian Federal Ministry of Health have set a mandate for universal coverage, which is defined as the provision of one LLIN for every two people at risk or one net per sleeping space with mass free distribution campaigns, typically on a three-year cycle [22]. While Ethiopia has made incremental progress in tool provision, historical data reveal a consistent “utilization gap” with suboptimal population-level utilization [23,24], as the functional survival time of nets in field settings is often shorter than the intended replacement cycle [17,25].

The epidemiological landscape of malaria in Ethiopia is currently defined by a concerning resurgence, with national incidence increasing by 32% between 2021 and 2022 and the total number of clinical cases doubling in 2023 compared to the previous year [4]. This reversal in progress is attributed to a complex interplay of factors, including vector behavioral adaptations toward early evening outdoor biting [26–28], insecticide resistance [29,30], and persistent socio-behavioral barriers to intervention compliance [13,31]. In 2023, the Ethiopian government launched a massive targeted distribution campaign, aiming to restore universal access across endemic regions to address these challenges [32]. Updated national evidence on the effectiveness of LLIN is particularly critical, including identifying the socio-behavioral and determinants of LLIN effectiveness to achieve Ethiopia’s 2030 goal of indigenous malaria elimination. A post-distribution survey was carried out to assess net ownership, usage, and condition to evaluate the effectiveness and appropriateness, including whether the distribution process was transparent, and whether there were any unintended negative impacts, including protection risks. Therefore, this study aimed to assess the coverage, accessibility, and utilization patterns of Long-Lasting Insecticidal Nets (LLINs) and to identify the socio-behavioral factors among households in malaria-endemic communities of Ethiopia. This approach will help bridge the gap between LLIN distribution and effective malaria prevention, ultimately informing strategies to improve the impact of malaria control programs across Ethiopia’s endemic regions.

## Materials and Methods

### Study Setting and Design

A community-based, cross-sectional survey was conducted from October 2024 to January 2025 to evaluate the coverage, access, and utilization of Long-Lasting Insecticidal Nets (LLINs) in Ethiopia. The study targeted malaria-endemic areas across 11 administrative regions, encompassing both urban and rural settings of Ethiopia (**Figure 1).**

**Figure 1:**
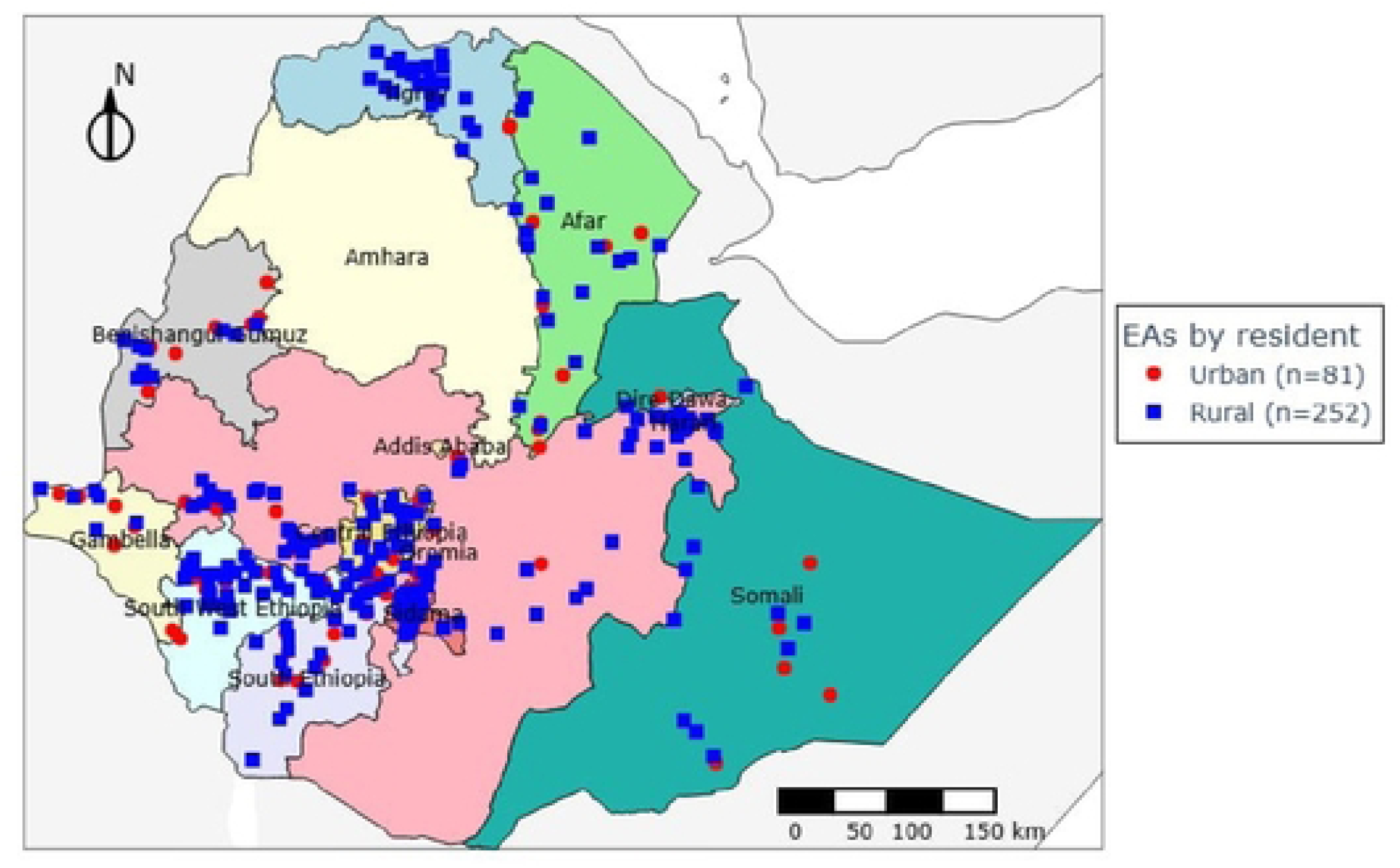
Enumeration areas (EAs) distribution across survey regions, Post LLINs distribution survey, 2024.

### Sampling Frame and Stratification

The sampling frame was constructed by integrating the 2019 Ethiopia Population and Housing Census data with updated LLIN distribution registries. The Primary Sampling Units (PSUs) were defined as Enumeration Areas (EAs), totaling 87,550 units nationwide. Stratification was achieved by cross-classifying the administrative regions by place of residence. A two-stage stratified cluster sampling technique was employed to ensure national representativeness. In the first stage, Enumeration Areas were selected using a Probability Proportional to Size (PPS) method based on the number of households per EA. In the second stage, a complete household listing was conducted in each selected EA, with segmentation applied to units of 30 households to maintain manageable data collection (**Figure 2**).

**Figure 2:**
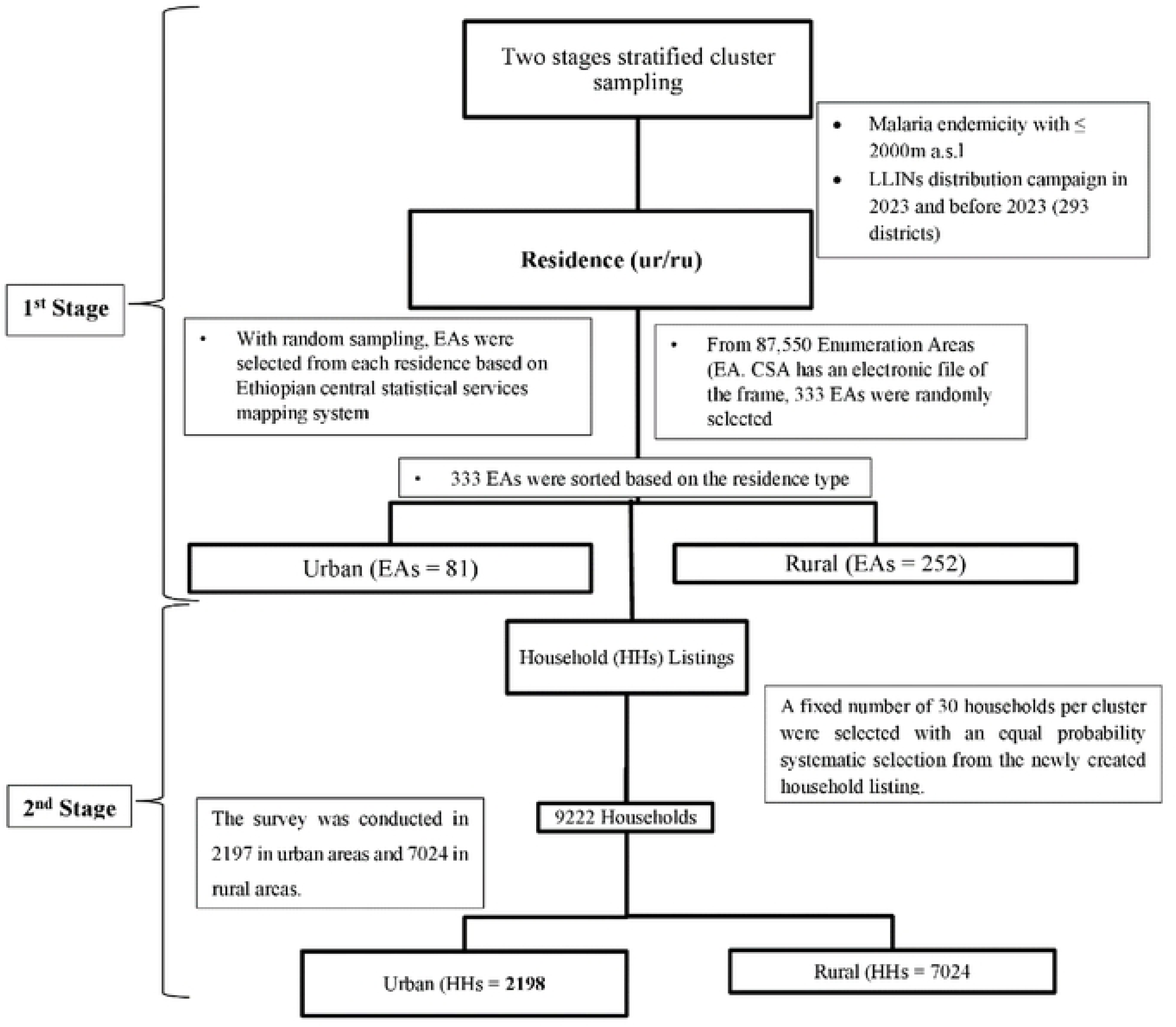
Systematic sampling procedures, Post LLINs distribution survey, 2024.

### Data Collection

Data were collected via face-to-face interviews and direct physical observation using standardized, structured questionnaires deployed through Open Data Kit (ODK) Collect. Household interviews were conducted with heads of households or eligible adults to obtain data on demographics, ownership, and usage patterns. Quality control was maintained through a preliminary pilot survey and daily supervisor validations to ensure data completeness before server transmission. During these visits, collectors performed direct physical observations to verify the presence, installation, and physical integrity of the nets. At the central level, data submitted to the dedicated server underwent real-time monitoring for outliers or clusters with unusual non-response rates. Any discrepancies identified during the synchronization process were immediately communicated back to the field teams for clarification or corrective action before the conclusion of the data collection period in each cluster.

### Management and Analysis

Data analysis was performed using SPSS version 28.0. To account for the complex survey design, the analysis utilized the SPSS Complex Samples module, incorporating cluster and strata variables to produce design-based standard errors and 95% confidence intervals. The primary outcome variable was defined as a binary indicator of LLIN utilization, specifically whether a respondent slept under an insecticidal net the night preceding the survey. To identify independent predictors of use, a hierarchical multivariable logistic regression was conducted with a random intercept at the household level to account for the clustering of individuals. Independent variables were entered in four conceptual blocks: sociodemographic factors, household factors, self-efficacy regarding net hanging, and knowledge or misconceptions about malaria.

Variables were retained in the final regression model based on a significance threshold of *p* < 0.05 in likelihood ratio tests. Regarding ethical standards, the study protocol was approved by the Institutional Review Board of the Ethiopian Public Health Institute (EPHI-IRB). In recognition of varying literacy rates, oral informed consent was obtained from all participants after explaining the study objectives and the voluntary nature of participation. The research was conducted in strict adherence to the Declaration of Helsinki, ensuring that all data remained confidential and were used exclusively for the purpose of this scientific investigation.

## Results

### Survey Population Characteristics

The post-net distribution survey was conducted in 11 regions of Ethiopia to assess the ownership and utilization of Long-Lasting Insecticide Nets (LLINs). The survey covered 9,222 households comprising 34,427 individuals. Of this population, 26,681(77.5%) individuals resided in rural households and 7,746 (22.5%) in urban households. Similarly, 76.2% of the households (7,024) were in rural areas, while 23.8% (2,198) were urban, indicating a clear rural predominance. The age distribution indicates a young population, with the 25–49 age group comprising the largest segment (37.12%,12,778 individuals). This was followed by children and adolescents aged 5–14 years (25.61%, 8,818) and young adults aged 15–24 years (18.81%, 6,476). Children under the age of five (0–4 years) represent 10.88% (3,747) of the total population. Across age groups, the majority (76% to 80%) of individuals live in rural settings, reflecting a uniform rural predominance. The survey population was balanced by gender, with males constituting 50.91% (17,526) and females 49.09% (16,901). Among the females surveyed, 1,626 were pregnant women, a group specifically targeted for malaria protection (**Table 1**).

### LLINs ownership and utilization

The current survey result has shown the overall net ownership of 71.5% (CI: 70.5%-72.4%), with a marked difference in ownership among regions, with Gambela (95.0%, 95% CI: 93.0-96.6), Afar (94.3, 95% CI: 92.5-95.7), and Benishangul Gumuz (90.0%, 87.2-92.5] demonstrated highest ownership. In contrast, regions such as Somale and Central Ethiopia showed low LLIN ownership of 32.7% (95% CI: 27.9-37.9) and 54.3%95% CI: [51.1-57.4), respectively. Furthermore, there was a notable variation in ownership between residential settings, with rural areas demonstrating higher ownership of 75.1% (95% CI: 74.1%-76.1%) compared to low ownership 24.9% (95% CI: 23.9-25.9) in urban areas. Moreover, even though there was variation in ownership between residential settings across regions, the result revealed a consistent trend of higher net ownership in rural areas than urban settings in all regions, except for Gambela, where net ownership was relatively comparable between rural and urban settings (**Table 2**). This 10-percentage point range reflects less precision in the estimate for this region compared to others, which may be due to greater variation in net distribution within that region. The implication of these intervals also highlights where the distribution is most equitable. In most regions, the urban and rural settings 95% CIs were distinct, reinforcing the rural focus of net distribution. For example, in Oromia, the urban CI (16.1-20.3%) and rural CI (79.7-83.9%) were widely separated. However, in Gambela, the urban CI (44.4-52.8%) and rural CI (47.2-55.6) showed significant overlap. This overlap implies that, within Gambela, there were no statistically significant differences in LLIN ownership between urban and rural households, representing the most balanced regional distribution recorded (**Table 2**).

Among a total surveyed household population of 34,427 individuals, 22,612 individuals living in 6588 households that owned at least one net had access to LLINs (Table 2). The adequacy of LLIN distribution was assessed using the WHO standard of one net for every two persons. This was calculated by multiplying the number of nets by two and dividing it by the total population within households of LLIN owners. The overall proportion of LLINs for every two persons at the national level was 58.3%, with the highest proportion in urban settings (60.8%) compared to rural settings (57.5% (Figure 2). The findings showed a marked variation in the adequacy of net supply relative to the population size across the surveyed regions. Somali and Afar regions reported the highest proportions of LLINs for every two persons at 94.1% and 92.2%, respectively. Hareri also showed a high distribution-to-person ratio at 82.8%. Conversely, several regions fall below the 50% threshold, meaning less than half of the population was covered by the “one net for every two people” standard. These lower-performing regions include South Ethiopia (48.4%), Central Ethiopia (49.6%), and Benishangul Gumuz (49.9%). Oromia, which has the largest surveyed population of 7,070 individuals, demonstrated a proportion of 54.8%, which is below the national average. Similarly, other regions with large populations, such as Sidama (4,425 people) and Southwest Ethiopia (4,398 people), reported proportions of 55.7% and 57.0%, respectively. In contrast, Gambela managed a proportion of 62.9% for its population of 1,838, exceeding the national average 58.3%, indicating a more robust distribution relative to its demographic size (**Figure 3**).

**Figure 3:**
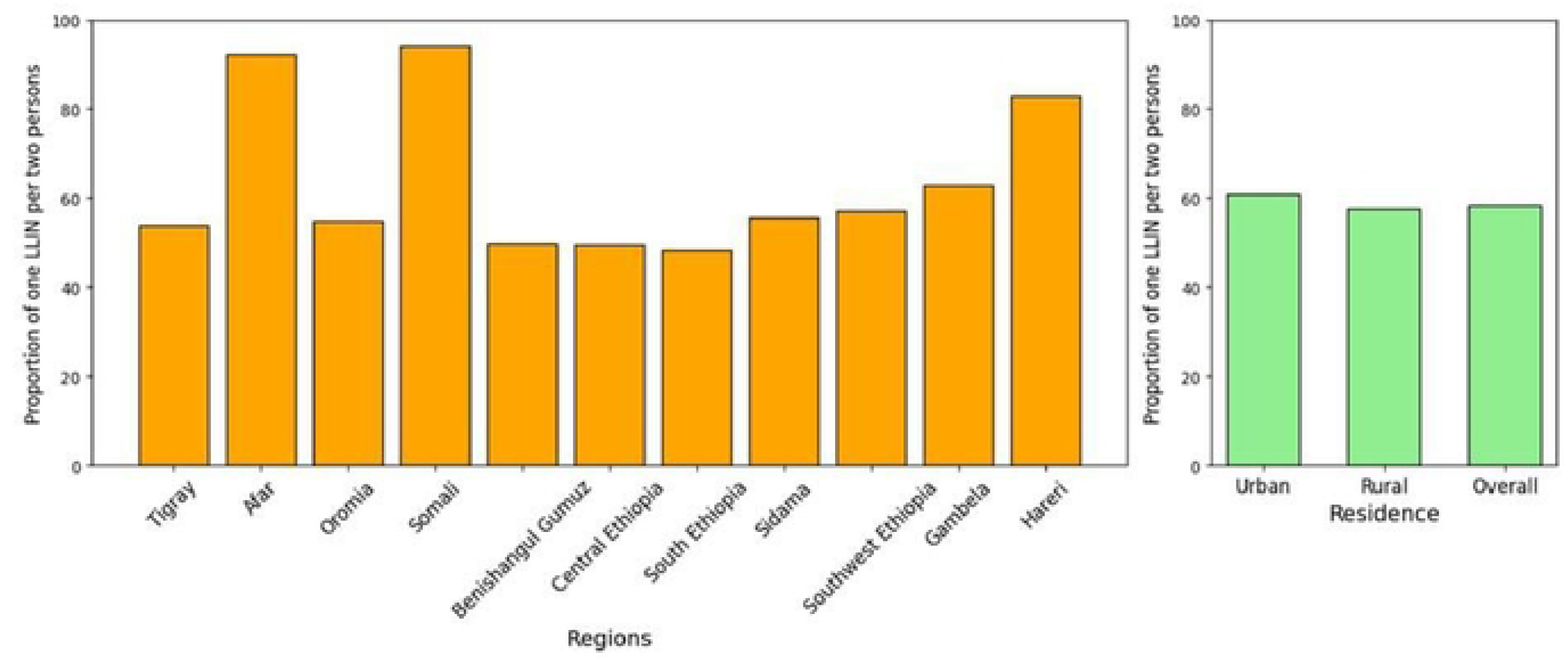
Proportion of insecticide-treated nets for every two persons across regions and residence type, post-net distribution survey, 2024.

The overall LLIN utilization among household members who owned at least one LLIN was 59.9%, representative of the surveyed population. Utilization varies by residence, with rural households accounting for 72.7% (95% CI: 71.9%–73.5%) and urban households 27.3% (95% CI: 26.5%–28.1%). Moreover, there was regional variation in LLIN utilization, with Gambela (94.3%, 95% CI: 93.2–95.4) and Afar (92.2%, 95% CI: 90.9–93.5) reporting the highest utilization, while Oromia had the lowest, 37.1% (95% CI: 35.7%–38.5%). Notably, the Somali region has a much wider 95% CI (47.2%–60.1%) compared to other regions (Table 2). In Tigray and Hareri, utilization proportion was recorded at 100% in rural settings, with no urban distribution and utilization reported in the sample. However, Gambela demonstrated similar utilization between urban (47.6%, 95% CI: 45.2%–50.1%) and rural (52.4%, 95% CI: 49.9%-54.8%) (**Table 3**).

We analyzed LLINs utilization among vulnerable groups, including pregnant women and children under five. Pregnant women achieved the highest LLIN utilization rate at 78.5% (846/1078; 95% CI 76.0%–80.0%). Among the general population, females reported higher utilization (61.9%; 95% CI: 61.0–62.8) than males (57.9%; 95% CI: 61.0–62.8). Utilization across age groups showed distinct patterns, with the highest use occurring among adults aged 25–49 (68.7; 95% CI: 67.7–69.7) and children aged 0–4 (67.2%; 95% CI: 65.3–69.0). In contrast, school-aged children (5–14 years) had the lowest utilization rate at 49.7% (95% CI: 48.4-51.0). Regression analysis confirmed these findings, with age (OR=0.258, 95% CI: 0.216-0.39; p<0.001) and pregnancy status (OR=0.66, 95% CI:0.594-0.738; p<0.001) being statistically significant predictors of LLIN use. Conversely, gender was not statistically significant (OR=0.985, 95% CI: 0.909-1.067; p = 0.704). Across all demographic categories, LLIN utilization was consistently higher in rural areas, accounting for approximately 72% to 79% of total net use. The most pronounced rural concentration was seen among adults 50+ age group (79.1%), while the most balanced urban/rural distribution among age groups was observed in the under-five (0–4 years), where 28.5% of utilization occurred in urban settings (Table 3). These narrow confidence intervals across the urban/rural split confirm that the rural-dominant trend of net utilization was consistent across every surveyed demographic (**Table 4**).

### Associated LLINs use and ownership barriers

In this survey, we assessed household criteria for mosquito net distribution, sleeping practice, and preference for net color and shape. The findings showed that 88.2% (95% CI: 87.4-89.0) of households reported mosquito net distribution was based on family size. In contrast, only a small fraction (3.6%, 95%CI: 3.1-4.0) believed sleeping space was the criterion, while 6.9% (95%CI: 6.3-7.6) did not know the criteria. Sleeping practice showed that most household members do not sleep outdoors (87.0%, 95% 86.3-87.7), whereas 13.0% (n = 1,198) reported sleeping outside. Among those who slept outdoors, 64.2% (95% CI: 61.4-66.9) reported using bed nets, while 35.8% (95% CI: 33.1-38.6) did not, leaving them vulnerable to malaria. The 95% CI for the outdoor net utilization (61.4%-66.9%) shows that while a majority seeks protection, there is still significant room for improvement in ensuring consistent net use during outdoor sleeping. Preferences for net color varied considerably. Blue was the most preferred color at 42.2% (95% CL: 41.2%-43.2%), followed by green (26.5%, 95% CI: 25.6-27.4), white (16.3%, 95% CI: 15.5-17.0) and 15% (95% CI: 14.3-15.7) expressed no preference Furthermore, Net shape preference showed a clear preference for rectangular nets, with 66.8% (95% CI: 65.8-67.7) of respondents choosing them over conical shape (21.7%, 95% CI: 20.8-22.5) (**Table 5**).

The current results indicated that a majority of respondents (77.2%, 95% CI: 6.4%-78.1%) had heard messages about mosquito nets, suggesting that basic awareness was widespread and consistently reported. However, direct home-based education reached fewer people, at 40.2%, while a majority (57.1%) reported they had not recently received such information at home. This suggests that while mass communication is reaching most people, personalized household-level education is less frequent. When information is delivered at home, Health Extension Workers were the primary source, accounting for 74.3% of the education provided followed by Health care workers at 19.7%, while other sources like community health workers (3.5%) and peer educators (0.2) played a much smaller role. Despite these education efforts, there was a clear practical skill gap, 60.5% of the respondents reported they were not confident in hanging a mosquito net, highlighting a critical area for future behavioral change interventions. The surveyed population generally holds positive views regarding the utility and safety of nets. Over 70% of respondents expressed high levels of confidence in the importance of children sleeping under a treated net (25.7% extremely confident and 44.5% very confident). Similarly, 73.5% were confident that treated nets are safe to sleep under. However, a notable misconception persists regarding malaria transmission: 64.1% of respondents agreed (strongly or somewhat) that people were only at risk during the rainy season. Even though 24.2% strongly agreed and 39.9% somewhat agreed about the risk of malaria during the rainy season, the findings indicated that people trust the nets; many may not recognize the need for year-round protection (**Table 6**).

Regarding LLINs maintenance, the survey findings showed that the population was nearly evenly split, with 51.1% of households (n = 4708) reporting that they washed their nets and 48.9 stating they did not. Among those who did wash their nets, the majority (67.5%) do so reactively when the net appears dirty, rather than on a fixed schedule. Regular washing was less common, with only 18.2% of respondents washing two to three times a year and only 7.9% washing once a year. Regarding the materials used for cleaning, bar soap was the most prevalent choice, used by 54.7% (95% CI: 53.2-56.1) of households. Detergents like OMO were the second most common cleaning agent at 32.2%, while 7.9% of respondents wash their nets using only water. Harsh chemicals like bleach were rarely used, reported by less than 1% of the respondents. Drying practices revealed a strong adherence to keeping nets out of direct sunlight, which was crucial for preserving the insecticide. A combined 94.5% of households reported drying their nets in the shaded areas, with the most common location being outside on the ground in the shade (45.3%), followed by a cloth line in the shade (25.2%), and inside a shaded area (18.4%). Only a small fraction of respondents (5.5%) reported drying their nets in direct sunshine, a practice that can degrade the net’s protective chemicals. The high percentage of shaded drying suggests that current public health messaging regarding proper net care was effectively adopted by the vast majority of those who maintain their nets (**Table 7**).

## Discussion

The 2024 post-LLIN distribution survey provides a comprehensive assessment of Ethiopia’s progress towards universal coverage and effective utilization of long-lasting insecticide nets (LLINs). National household LLIN ownership reached 71.5%, a modest increase from 64% in 2015 [23], 64.8% in 2017 [33], and 67% in 2020 [24]. Despite this upward trend, coverage remains below both the national target of 100% for malaria endemic households [32] and the WHO benchmark of 80% universal coverage [4], indicating the current LLIN distribution is insufficient to meet global elimination milestones [20,23]. Additionally, our survey revealed that LLIN access, defined as one net per two people or one net per sleeping space for all households in malaria-endemic areas, improved to 58.3% compared to the reports of 32.0% in 2015 [23,33], 39.3% in 2017 [33], and 40.3% in 2020 [24]. This persistent gap indicates that while mass distribution campaigns have successfully increased net numbers across years, they have not yet achieved the optimal adequacy required to protect all household members. Historically, approximately 81% of nets have been distributed in rural areas, reflecting the higher transmission risk in rural, low-altitude settings [33].

We also observed urban-rural differences in LLIN ownership. In the current survey, urban households have shown slightly higher coverage than rural households. This finding contrasts with earlier national surveys, where rural households consistently have higher ownership than urban households [23,33]. Despite these shifts, both settings have shown improvement over time, yet neither has achieved the threshold required for universal protection. These findings suggest that distribution campaigns have expanded coverage, but geographic inequities remain a persistent barrier to equitable malaria prevention. Relatively higher urban adequacy likely reflects smaller average household sizes in urban settings, as protection levels typically increase as family size decreases because fewer nets are needed to meet the WHO 1:2 ratios [24]. Historically, ownership has been higher in rural areas (70.0%) compared to urban (55.4%) settings in 2020 (24) because the rural population faces the highest transmission risks due to environmental factors like irrigation and proximity to water bodies [34].

The high ownership rates in Gambela (95.0%) and Afar (94.3%) align with the 2020 national survey, which also identified these regions as leaders in net ownership with 85.8% and 82.6%, respectively [24], attributed to their status as a high-transmission western and eastern rift valley lowland region [32]. Notably, Gambela was the exception where urban and rural ownership overlapped, indicating a highly balanced and equitable distribution system similar to previously reported findings, with better-than-average community awareness and access [24,33]. Conversely, the low ownership recorded in the Somale region (32.7%) represents a sharp decline from the 63.7% reported in 2017 [33] and 65.3% reported in 2020 [24].

In contrast, Oromia (54.8%), which is falling below the national average, underscores the difficulty of maintaining universal coverage, which is most pronounced and is supported by research showing heterogeneity in access and utilization in Arsi zones [35,36]. The adequacy proportion of 54.8% in this region has shown an improvement compared with the 31.0% of adequacy that was observed in 2015, but it remains below the current national average, likely due to the logistical complexities of serving its large population [23,33]. Similar to Oromia, the recorded ownership in Sidama (55.7%) has fallen below the national average [24,37] but is considerably higher than recent longitudinal reports from the region, which found adequacy dropping from 36% to 29% in late 2023 [25].

The 94.1% and 92.2% adequacy proportions (proportion of one LLIN for two persons) reported for the Somali and Afar regions, respectively, mark a dramatic shift in LLIN adequacy relative to population size. In 2015, Somali households achieved a 1:2 ratio of only 30.7%, while Afar was recorded at 28.4% [23]. These regions have transitioned from being below the national average to exceeding international targets, contrasting with previous results where they were among the lower-performing strata [23,33]. Several regions: South Ethiopia (48.4%), Central Ethiopia (49.6%), and Benishangul Gumuz (49.9%) demonstrate a LLIN shortage, with less than half of the population covered by the universal standard, and need to be prioritized for future distribution efforts [33]. Consequently, sustaining these high distribution-to-person ratios will require not only frequent replenishment but also intensified social and behavior change programs to ensure available supply is utilized consistently and maintained effectively [25,33]. Scientific evidence suggests that the impact of these distribution efforts is fundamentally constrained by rapid net attrition and physical decay, with a median functional survival time of only 19 months, meaning half of the distributed supply becomes unserviceable well before the standard three-year replacement cycle [17]. Consequently, achieving malaria elimination will require a strategic transition from commodity-centric distribution toward intensified Social and Behavior Change (SBC) programs that focus on practical skill-based education and personalized communication to ensure consistent, year-round utilization [21,33,38].

The current post LLINs distribution survey reported that the overall LLIN utilization of 59.9% among household members who owned nets represents a substantial improvement in behavioral adoption compared to historical national utilization. This utilization level is higher than the 40% reported in 2015 [23], the 42.6% in 2017, and the 44.2% identified in the 2020 national survey [33]. However, despite these gains, utilization remains below the conventional 80% threshold required to provide sufficient community-wide protection [25,33]. The disparity between residential settings, where rural areas account for 72.7% of total utilization, and the strategic rural focus of Ethiopia’s malaria control program. This trend of higher rural utilization contrasts with the 2020 national survey, which found more equitable usage between urban (47.4%) and rural (43.4%) households [24]. Historical data from 2007 and 2017 indicated that urban areas sometimes possessed better access to health information and higher use rates among specific groups, a pattern that the current results suggest may be shifting back toward rural dominance (24,33,35). However, this finding contrasts with earlier studies and the 2007 Malaria Indicator Survey, which suggested that urban areas sometimes had better access to information and higher use rates among specific vulnerable groups [33,35].

Regional variations in the current survey further highlight persistent gaps. The high utilization rates in Gambela (94.3%) and Afar (92.2%) were consistently reported to have the highest national coverage and usage, aligning with findings from the 2017 and 2020 LLIN surveys [24,33]. Conversely, Oromia’s utilization of 37.1% is a critical outlier, identifying it as a high-priority area for future social and behavior change communication (SBCC). Previous research in the Kersa and Miesso district of Oromia regional state also found that rural residents were significantly more likely to utilize LLINs than their urban counterparts [39,40]. This variation is consistent with reports that local factors, such as mosquito abundance and perceived risk, significantly influence net use [38]. The higher variability in net use, such as in the Somali region, is possibly due to the migratory nature of pastoralist populations or logistical challenges in distribution [25,33]. The data on equity and setting-specific proportions underscore the importance of local context in malaria elimination. For instance, Gambela stands out as the most equitable region, with net use nearly identical between its urban and rural populations. This achievement suggests that distribution strategies and health education have been uniformly effective across different residential environments in the region. The pregnant women’s LLIN utilization 78.5%) and children aged 0–4 years at 67.2% among vulnerable groups were a marked increase from 2015 national figures of 44% for pregnant women and 45% for children under five [23,24,33]. Despite these achievements, a distinct age-related “utilization gap” was identified in school-aged children 5–14 years), the group least likely to be protected [37]. This is particularly concerning given that age and pregnancy status remain the most powerful predictors of LLIN use [35,37].

The low utilization progress in general is recorded below the national and international threshold, which may be due to the identified behavioral and logistical factors like outdoor sleeping habits and net design preferences, providing critical insights into malaria prevention and control in Ethiopia. The family size (88.2%) was recognized as the primary factor for determining the number of nets received, which reflects a high level of community awareness and alignment with national policy, mirrors the Ethiopian National Malaria Guidelines, which mandate distribution based on the number of household residents, specifically, one net for every two people [25,32,41]. This understanding was a marked improvement over historical challenges in distribution logic, where some regional managers noted that a lack of precise quantification by residents often led to over- or under-distribution [25]. Contrastingly, in other international contexts like the Dominican Republic, quantification often relies on assumed sleeping spaces rather than resident counts, which has led to significant coverage gaps [6]. The reported 13.0% of individuals sleeping outdoors represents a significantly vulnerable population, particularly as vectors in some regions have shown behavioral adaptations toward frequent outdoor biting earlier in the evening [42]. While the 64.2% utilization among outdoor sleepers indicates a strong desire for protection, the 35.8% who do not use nets are at high risk. Scientific literature confirms that the perceived low density of mosquitoes and the discomfort of heat are primary barriers to net use in outdoor or non-conventional sleeping arrangements [7,42], and difficulties in hanging and replacing nets are primary factors contributing to suboptimal use [7]. This practical barrier is particularly problematic given that LLINs in Ethiopia have a median functional survival time of only 19 months, meaning nets are often damaged or discarded well before the end of the three-year distribution cycle [17]. Without the technical skills to properly mount nets, especially in rooms that serve multiple functions during the day, household members are more likely to leave nets unhanged or stored away [21]. This suggests that this is a stable behavioral trend that requires targeted Social and Behavior Change (SBC) programs to encourage consistent net use during outdoor activities and sleeping [7,25]. The clear preference for blue nets (42.2%) and rectangular shapes (66.8%) provides actionable data for procurement and distribution, aligned with the 2020 Ethiopian national survey, which reported a massive proportion of 99% preference for rectangular nets and a 72.2% preference for the color blue [24]. Some other studies in Ethiopia also suggested that conical nets might be preferred in certain settings due to ease of hanging in smaller rooms [21]. Considering these user preferences is essential for preventing repurposing and avoidance, as individuals are more likely to use nets they find attractive and convenient for their specific sleeping arrangements [21,42].

The surveyed population generally holds positive views on net safety or handling (73.5%), and the importance of protecting children, yet these attitudes are undermined by a fundamental misconception regarding transmission. The finding that 64.1% of respondents believe malaria risk is limited to the rainy season is a major obstacle to elimination goals. This seasonal perception is a leading reason for non-use in Southern Ethiopia, where nearly 30% of households report not using nets during the dry season because they believe there is “currently no malaria.” [38]. This behavior is often driven by a perceived low density of mosquitoes and physical discomfort from heat during dry periods [7,42]. To achieve national elimination, social and behavior change (SBC) programs must move beyond basic awareness to address these specific skill gaps and the critical need for consistent, year-round utilization across all climatic settings [24,38].

The reported awareness regarding mosquito net messages reflects a widespread reach of basic malaria information, showing a slight improvement over the awareness recorded in the 2015 MIS [23]. This finding is relatively encouraging, with the finding that only 40.2% received home-based education suggesting a persistent gap in personalized community outreach. Historically, Health Extension Workers (HEWs) have been the backbone of Ethiopia’s malaria strategy, responsible for approximately 81% of net distributions and the majority of community-level education [24]. The previous survey finding, HEWs were the primary information source (74.3%) of national data, aligned with where they reached over 92% of households in health messaging [24]. However, previous national assessments found that while coverage is high, only 33.7% to 38% of households could accurately recall key mosquito net messages, indicating that mass communication often fails to achieve the necessary behavioral depth [24,33]. The finding that 51.1% of households reported washing their nets indicates a stable behavioral trend compared to the 49% washing rate documented in the previous survey [24]. The reactive approach to maintenance, where 67.5% of households wash nets only when they appear dirty, mirrors the 2015 Ethiopia National Malaria Indicator Survey [23], where “when it gets dirty” was the primary trigger for washing across both urban and rural settings. Such behavior is programmatically significant because the frequency and method of washing are major determinants of how long a net retains its insecticidal bio-efficacy and physical integrity [6,21]. Regarding washing materials, the preference for bar soap (54.7%) is considered an adequate practice as it was less damaging than harsher alternatives. Conversely, the 32.2% use of detergents such as OMO identifies a critical area for social and behavioral change interventions, as these are classified as aggressive products that can rapidly reduce the net’s insecticidal efficacy [6]. Nevertheless, the current results show significantly higher adherence to safe materials than in other malaria foci; for instance, in the Dominican Republic, between 64.6% and 85.8% of households used aggressive products like bleach or chlorine [6]. The rarity of bleach use (less than 1%) in the surveyed population further suggests that Ethiopian households generally avoid the most chemically destructive maintenance practices compared to global trends [6]. A major success revealed by the survey is the 94.5% adherence to shaded drying, which is essential for protecting the insecticide from UV degradation [6,17]. This finding represents a stark contrast to other field settings where 75% to 90.5% of households dry their nets in direct sunlight [6]. The high percentage of shaded drying and the low proportion of individuals drying nets in direct sunshine (5.5%) imply that public health messaging and community education regarding proper net care have been effectively adopted by the vast majority of net owners [17]. Sustaining these maintenance behaviors is critical for extending the median functional survival time of LLINs, which in Ethiopia is currently estimated at only 19 months [17].

### Conclusion

While national ownership is improving, the persistence of regional disparities and the rapid attrition of nets often lasting less than the expected three years suggests that increasing coverage alone is insufficient. Continuous distribution mechanisms, such as routine delivery through antenatal care and immunization clinics, remain essential to sustain gains and address the “loss” of LLINs between mass campaigns. While national adequacy is rising, the “net use gap” remains primarily driven by insufficient access within the household rather than a failure of behavioral compliance. National strategies have successfully ensured that populations with the highest vulnerability (pregnant women and children under 5) are prioritized for protection. However, internal household resource allocation patterns create a secondary protection gap for other demographic segments, such as school-aged children, who are frequently displaced from protected sleeping spaces as families prioritize younger siblings. A significant constraint to sustained efficacy is the disconnect between ownership and the practical knowledge required for proper installation and maintenance, identifying a technical barrier that limits the conversion of access into consistent use. This is further compounded by misconceptions regarding the seasonal nature of disease transmission, leading many to perceive protection as unnecessary during dry periods and thereby maintaining reservoirs for future transmission. While community adherence to certain maintenance behaviors reflects successful health education, these behavioral gains are often undermined by a reactive approach to care and the use of chemically aggressive washing agents that accelerate the loss of insecticidal efficacy. Additionally, residential disparities highlight the critical role of localized health extension networks, as rural environments demonstrate higher adoption rates compared to urban settings. While significant strides have been made in increasing availability, the simple provision of nets is insufficient to achieve the threshold necessary for community-wide protection and the interruption of transmission cycles. Ultimately, achieving malaria elimination requires a strategic transition toward skill-based interventions and personalized social and behavior change communication. Addressing demographic disparities, enhancing community engagement, and implementing continuous monitoring will be essential to bridge the gap between ownership and consistent, year-round protection.

## Acknowledgements

The successful completion of this national assessment was made possible through the intensive collaborative efforts of several key institutions. We extend our profound gratitude to the Ministry of Health (MoH), NIH grant D43 TW001505, the Regional Health Bureau, Zonal Health Department, and the District malaria focal point for their technical and logistical support. The data collection process relied heavily on the tireless efforts of field personnel and local health networks. We express deep thanks to the Health Extension Workers (HEWs), who serve as the backbone of the national malaria strategy and act as the primary information source for the majority of the surveyed communities. Most importantly, we are profoundly grateful to the 9,222 households and 34,427 individuals who participated in this survey. Their willingness to provide oral informed consent and share information regarding their domestic practices provided the critical evidence needed to evaluate current malaria control interventions.

## Data availability

The authors confirm that the data supporting the findings of this study are available within the article and its additional file. Raw data that support the findings of this study are available from the corresponding author upon reasonable request.

## Author Contribution

AW, GT, DY, HT., AE, and AG – research design and conceptualization. AW, AH, BG, EK, FG, SK, SY, and HTe – sampling frame and implementation. AW, HS., GA, DD, TK, SB, YW, BA., ST, and SH – regional coordination and strategic alignment with the National Malaria Strategic Plan. AW, GG, FT, and AE. – field supervision and data quality management. AW and SY– data analysis and writing of the original draft. GTr., MH., and YG. – validation, institutional oversight, and adherence to ethical standards. All the authors have read and agreed to publish the manuscript.

